# Emergency department admissions during COVID-19: explainable machine learning to characterise data drift and detect emergent health risks

**DOI:** 10.1101/2021.05.27.21257713

**Authors:** Christopher Duckworth, Francis P. Chmiel, Dan K. Burns, Zlatko D. Zlatev, Neil M. White, Thomas W. V. Daniels, Michael Kiuber, Michael J. Boniface

## Abstract

Supervised machine learning algorithms deployed in acute healthcare settings use data describing historical episodes to predict clinical outcomes. Clinical settings are dynamic environments and the underlying data distributions characterising episodes can change with time (a phenomenon known as data drift), and so can the relationship between episode characteristics and associated clinical outcomes (so-called, concept drift). We demonstrate how explainable machine learning can be used to monitor data drift in a predictive model deployed within a hospital emergency department. We use the COVID-19 pandemic as an exemplar cause of data drift, which has brought a severe change in operational circumstances. We present a machine learning classifier trained using (pre-COVID-19) data, to identify patients at high risk of admission to hospital during an emergency department attendance. We evaluate our model’s performance on attendances occurring pre-pandemic (AUROC 0.856 95%CI [0.852, 0.859]) and during the COVID-19 pandemic (AUROC 0.826 95%CI [0.814, 0.837]). We demonstrate two benefits of explainable machine learning (SHAP) for models deployed in healthcare settings: (1) By tracking the variation in a feature’s SHAP value relative to its global importance, a complimentary measure of data drift is found which highlights the need to retrain a predictive model. (2) By observing the relative changes in feature importance emergent health risks can be identified.

## Introduction

Predictive algorithms deployed in dynamic environments are subject to data drift, defined as a systematic shift in the underlying distribution of input features (i.e. *P*_*t*_(*x*) ≠ *P*_*t*+*t*′_(*x*), for probability distribution *P* defined at time *t*) ^1, 2^. In a clinical setting data drift occurs over time because of changing characteristics of presenting patients. In the long-term data drift can occur because of changes to national health policy (e.g., decreases in smoking rates^3^) and in the short-term from emergent diseases, most notably seen in the COVID-19 pandemic. For machine learning models, used as decision support tools, data drift can invalidate a model, as the drifted feature distribution may predominantly lie in a region where the model poorly performs. Due to the risk to patient care in using machine learning clinical risk scores, proactive model monitoring, and, model retraining is crucial to ensure they retain their high-performance. Machine learning models are also subject to concept drift, where the underlying statistical relationship between input features and target variables fundamentally changes (i.e. *P*_*t*_(*y*|*x*) ≠ *P*_*t*+*t*′_(*y*|*x*)) ^4, 5^. In clinical settings this could surmount to changes in operational procedure which now leads to different outcomes for patients presenting in similar ways to historical episodes. Assuring model performance in medicine is multifaceted, and requires both performance tracking and attribution of what is changing in the input features.

A prime example of data drift is the emergence of the COVID-19 pandemic, which has brought a severe, abrupt, and, continuous shift in circumstances across industries ranging from financial services to healthcare. In particular, the pandemic has led to an unprecedented change to patient landscape in hospitals, leading to fundamental restructuring of operations^6,7^. In the UK, the National Health Service (NHS) reported a 57% drop in emergency department (ED) attendances in April 2020, equivalent to 120,000 fewer attendances than the previous year^8^. The exponential increase in COVID-19 related attendances through the first wave (March-May 2020), matched by a sudden drop in acute visits for stroke and heart attacks required sudden and large changes to operational procedures in EDs^8^.

A key step in an ED’s operations is triage where patients are sorted by acuity, to identify those in most need of immediate medical intervention. Triage assesses a patient’s short term risk and potential care trajectory based on vital signs, demographics, chief complaints, and, condition history. Models which predict patient outcomes (e.g. whether a patient is likely to be discharged or admitted to hospital) seek to improve logistics in EDs and organise downstream resource allocation^9,10^. Such models could be particularly useful during periods of increased attendance, providing decision support when clinicians may be presented with a wealth of information without enough time to interpret and act upon it^11^.

We introduce a model (trained using attendances occurring prior to the outbreak of COVID-19) making use of basic, digitally available, patient information (e.g. age, arrival mode, chief complaint, vital signs) to predict whether a given patient will be admitted to hospital. Using the COVID-19 pandemic as an exemplar of data drift, we evaluate our model’s performance independently on both pre-pandemic and during pandemic patient cohorts, and use explainable machine learning to track data drift. We discuss how changes in model explanations and performance metrics together could be used as a complimentary trigger for identifying changing patient attendance behaviour, changes to ED operations, and identification of emergent diseases. For our admissions model, this enables an understanding of the most important factors of admission risk pre and during COVID-19, and attribution of why model performance drops during COVID-19.

## Methods

### Data

We used a (pseudonymised) patient attendance record of all adults to Southampton General Hospital’s ED (University Hospital Southampton NHS Foundation Trust - UHS) occurring from the 1st April 2019 to the 30th of April 2020. We considered only attendances resulting in admission or discharge (i.e. removing planned visits or referrals from other EDs), of which there are 82,402. To characterise each attendance for input into a machine learning classifier we included encoded patient descriptors (e.g., age), recorded chief complaints at registration and triage, attendance characteristics such as arrival time and arrival mode, and any vital signs recorded at the point of triage. All numerical variables were used directly as input to the model, whereas categorical variables are target encoded^12^.

We provided the classifier with a view of a patient’s medical history by making use of the historical discharge summaries associated with the patient, both from the ED and from the patients electronic health record. For every patient, all previously listed conditions at discharge were one-hot encoded to indicate if a patient had a given condition (e.g. hypertension, type 2 diabetes) recorded at a previous attendance. Additional discharge notes (e.g. lives alone, history of alcohol abuse, current smoker) were also included and encoded in this manner. The full list of input features provided to our classifier is given in Supplementary Table 1. The electronic health records used by our models are available to review by clinicians and are used in regular practice. Our model did not have access to any free text fields in the electronic health records. We split our data temporally into training (40,309 attendances), pre COVID-19 test (37,367 attendances) and COVID-19 test (4726 attendances) cohorts, as detailed in Figure 1.

**Figure 1.**
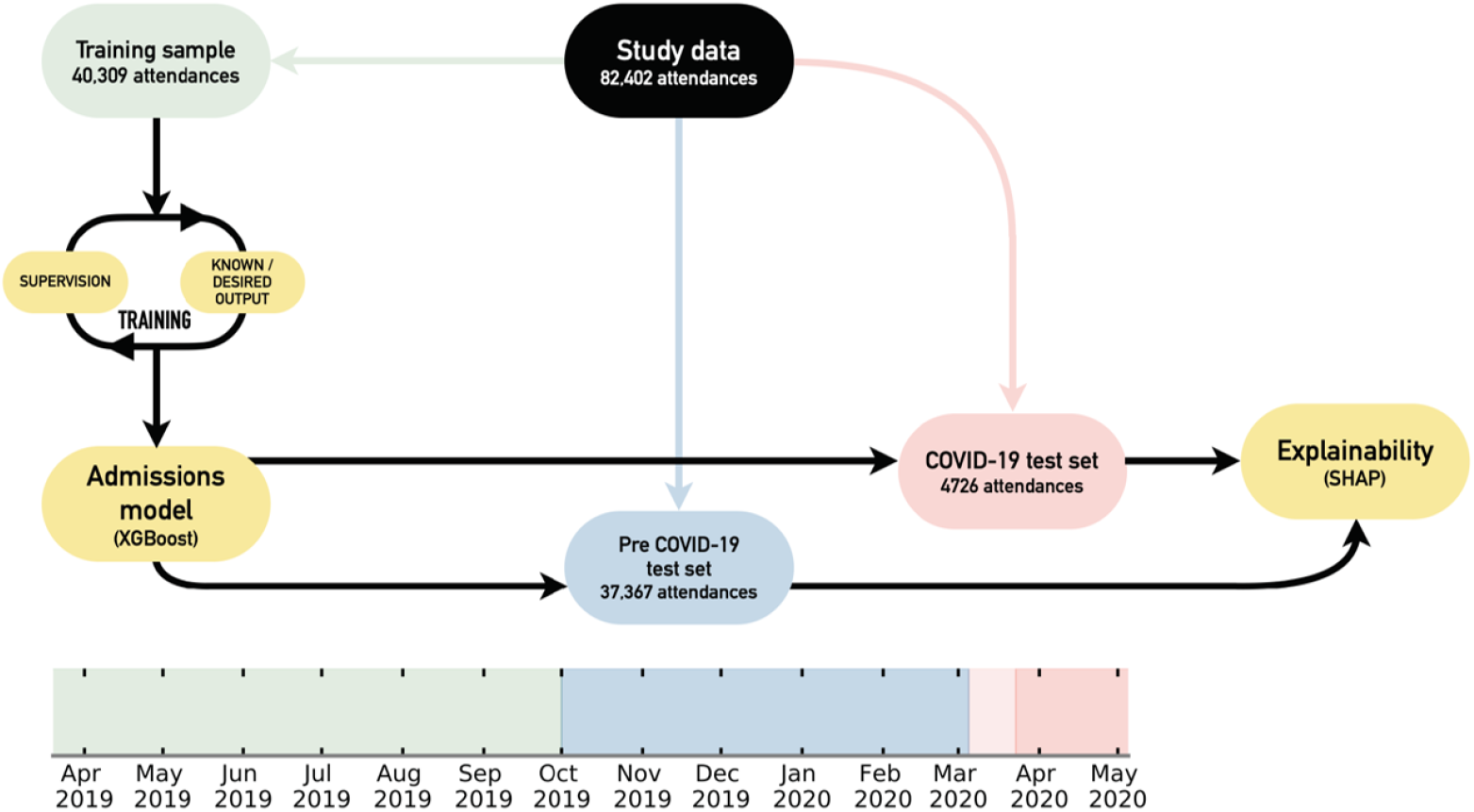
Schematic of study design. The emergency department admissions data is segregated temporally into training (green), pre COVID-19 test (blue), and COVID-19 test (red) cohorts according to the timeline. A XGBoost admissions model is trained and then tested to understand relative performance pre and during the COVID-19 pandemic. A SHAP explainability model is used for both test sets to investigate data drift. The darker red shaded region indicates when the UK entered a national lockdown (23rd March 2020), and, the start of the lighter shaded region shows the time between the first registered COVID-19 death in the UK and the national lockdown (included in pre COVID-19 test set).

### Ethics and data governance

This study was approved by the University of Southampton’s Ethics and Research governance committee (ERGO/FEPS/53164) and approval was obtained from the Health Research Authority (20/HRA/1102). All methods were carried out in accordance with relevant guidelines and regulations associated with the Health Research Authority, and NHS Digital. Research was limited to use of previously collected, non-identifiable information. Informed consent was waived by the University of Southampton’s Ethics and Research governance committee, University of Southampton, University Road, Southampton, SO17 1BJ, United Kingdom and the Health Research Authority, 2 Redman Place, Stratford, London, E20 1JQ, UK. Data was pseudonymised (and where appropriate linked) before being passed to the research team. The research team did not have access to the pseudonymisation key.

### Admission modelling

The aim of our admissions model was to predict patient outcomes (admitted or discharged) at the point of triage during an ED attendance. Predicting patient outcomes at this point in a patient’s care trajectory enables automation within the triaging system of the ED itself, ahead-of-time resource prediction, and downstream logistical planning. Admission models could also be used as part of a decision support system within the ED. Identifying patients at highest risk of admission, provides a sanity check for triage and helps ensure high acuity patients have timely medical intervention.

For our predictive algorithm, we used gradient boosted decision trees as implemented in the XGBoost framework^13^. XGBoost naturally deals with continuous, binary/discrete, and, missing data consistently; all of which are represented in our dataset. Model hyperparameters were selected using five-fold cross-validation of the complete training set using a sampler (Tree-structured Parzen Estimator) implemented with the Optuna library^14^. Model performance was evaluated independently by both test sets (pre and during COVID-19) using the Area Under the Receiver Operating Curve (AUROC) and with the precision recall curve. We note, that due to there being a significant shift in admission rate between test samples (panel b Figure 2), and the area under the precision recall curve is an inherently biased metric (i.e. depends on the class distribution), and, we further discuss its role in data drift interpretation in the results section.

**Figure 2.**
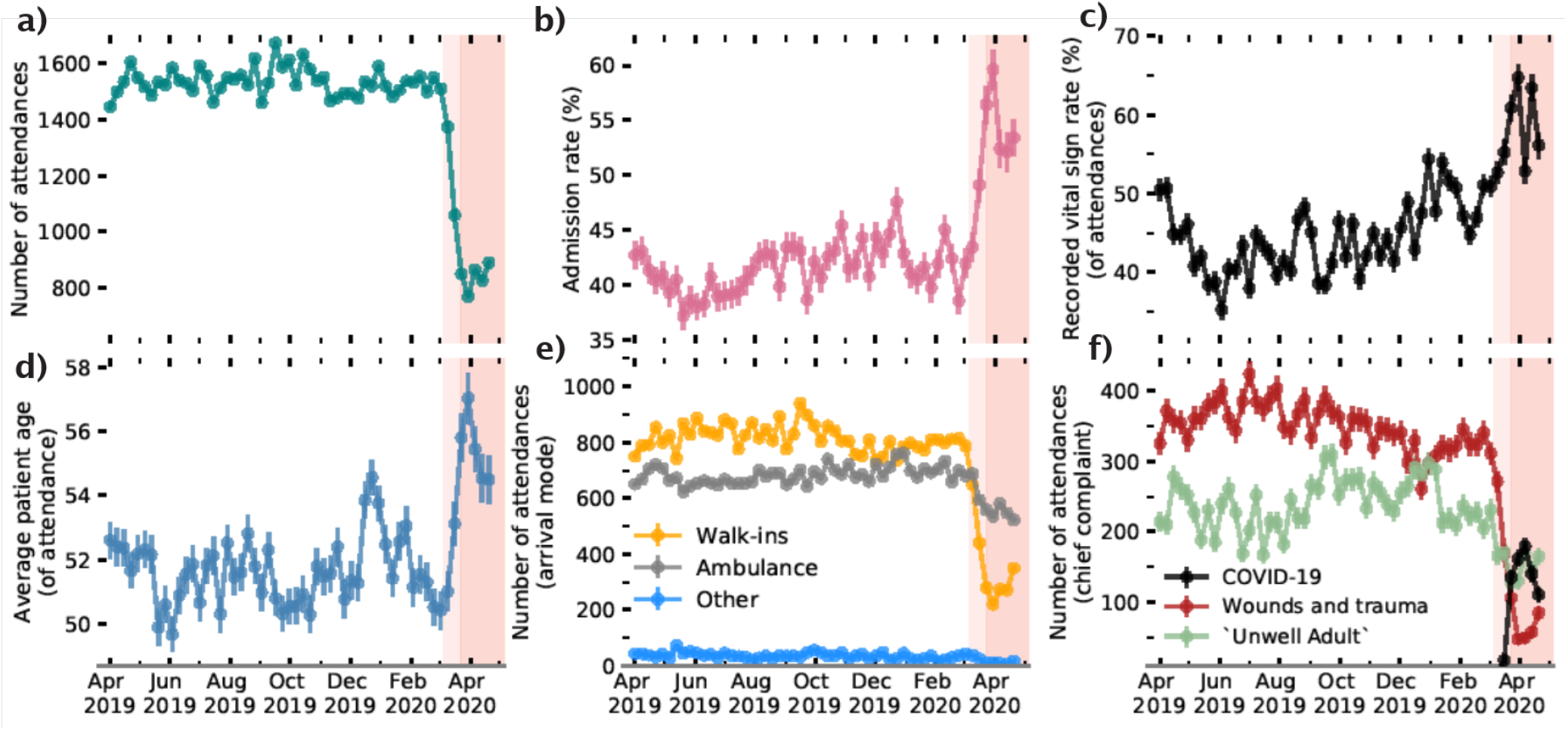
ED attendance statistics as a function of time. (Top row left-to-right) Number of weekly attendances, percentage of attendances that resulted in admission to hospital, and the percentage of attendances with vital sign measurements recorded. (Bottom row) Average age of the patient attending, mode of arrival to ED and chief complaints on attendance. Note: COVID-19 recorded at the point of discharge. The red shading is consistent with the timeline in Figure 1.

### Model explainability

To attribute the fractional contribution of a given input feature to admission risk, we made use of the TreeExplainer algorithm as implemented in the SHAP (SHapley Additive exPlanations) library^15–17^. TreeExplainer efficiently calculates so called Shapley (SHAP) values^18^, a game theoretic approach for attributing payout between coalitional players of a game. In the context of machine learning, SHAP values amount to the marginal contribution (i.e. change to the model prediction) of a feature amongst all possible coalitions (i.e. combinations of features).

There is a rich history of global interpretation for tree models which summarise the overall impact of input features on predictions as a whole. In a clinical setting, however, assurances must be made that all model predictions are interpretable, and every patient is evaluated fairly. One particular strength of Shapley values is that they are *locally accurate*, i.e, they can explain a prediction for a single attendance. Practically this means that the impact of each feature, per attendance, is defined as the change in the expected value of the model’s output when a feature holds this value vs. if it was unknown.

Feature importance can therefore be checked periodically by averaging over a fixed time period. Owing to their local accuracy, however, the sum of SHAP values for a given patient attendance must equal the patient’s admission risk predicted by the model. As a result, if the target distribution changes for a given period, the mean SHAP sum across all features will also change. Practically, since the admission rate increases during COVID-19, the mean magnitude of SHAP values increases accordingly. Therefore, to make temporal comparisons of relative feature importances we must normalise the SHAP values for a given attendance. To track data drift, we instead used absolute normalised (i.e. per attendance across all features) SHAP values. Normalizing the SHAP values (enforcing the sum of the SHAP values for a given attendance to equal one) in such a way makes the sum of the SHAP values of a given attendance invariant to changes in the target distribution.

## Results

### Attendance characteristics

A summary of weekly ED attendances for UHS throughout our dataset is provided in Figure 2. Specifically the onset of the COVID-19 pandemic led to a drop in ED attendances which led to sudden uptick in admission rate, average age of patients attending, and the fraction of attendances with recorded vital signs (see Figure 2). The drop of attendances appear to be driven by patients no longer ‘walking-in’ into the ED and because of a reduction in the number of attendances attributed to ‘wounds’ or related traumas (i.e. head injuries or limb problems). This is reflective of national attendance behaviour changes in the UK through the onset of the pandemic^8^.

### Detecting data drift

Implementing an admissions model within a dynamic environment such as an ED would require proactive monitoring to detect data drift. A popular choice to evaluate whether a model may need to be retrained is to track performance metrics over time^5^. In Figure 3, we show the performance of our trained model across our test data in weekly bins. The model’s performance (area under the receiver operating curve - AUROC) is shown for each week with bootstrapped 95% confidence intervals. The model remained stable until the onset of the pandemic. During the COVID-19 pandemic, there is a statistically significant (albeit small) drop in model performance. In the first few weeks of the pandemic (March 2020), however, there is no statistical degradation (i.e. average within the error bars), despite there already being significant evidence for changes to patient behaviour, and, ED operations (see Figure 2). It is difficult to ascertain whether the input features have shifted to be more closely distributed to regions where the model’s local performance is lower, or if this is a result of concept drift (i.e. change in the relationship between features and target variable which invalidates the learnt relationship).

**Figure 3.**
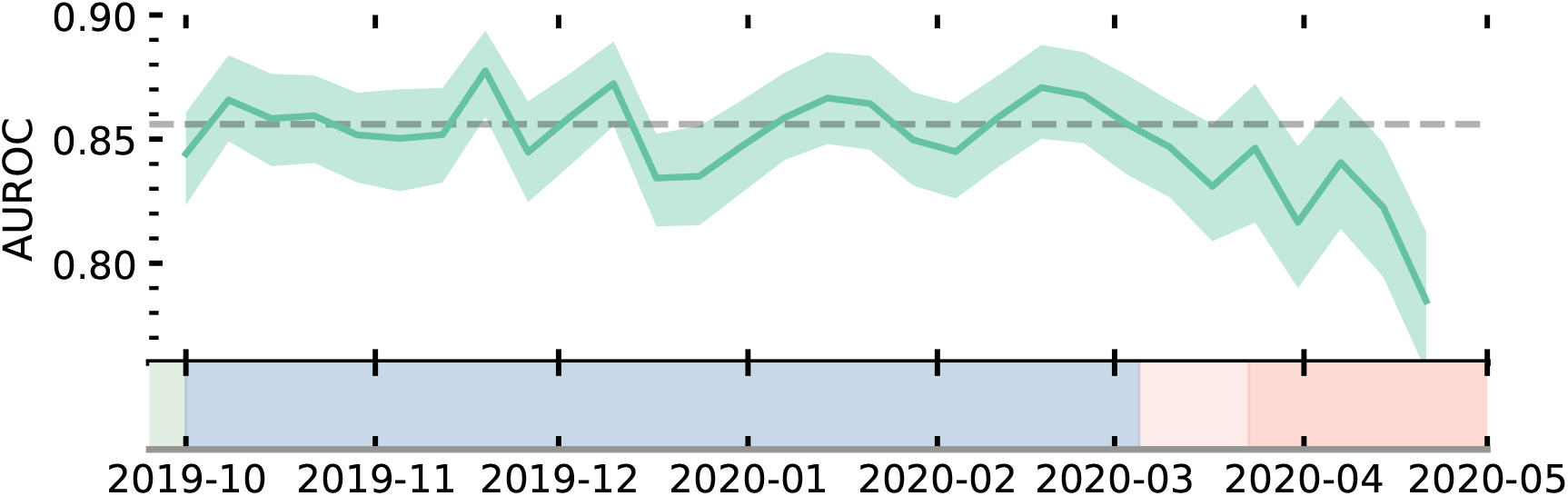
Time evolution of model performance evaluated by the area under the receiver operating characteristic curve (AUROC). AUROC values for the model are evaluated in weekly bins throughout the complete test period. Errorbars are 95% CI found from bootstrapping of each bin. The grey dotted line shows the average AUROC of the model evaluated on attendances occurring before the COVID-19 pandemic. Shading refers to the timeline defined in Figure 1.

The nuance of understanding whether retraining may be required is also highlighted when we consider the overall model performance for both our pre COVID-19 and during COVID-19 patient cohorts. In Figure 4, we show the ROC curve, precision-recall curve, and, a confusion matrix for both our pre COVID-19 (top row) and during COVID-19 (bottom row) test samples. Again, we see a statistically significant drop in the AUROC during the COVID-19 pandemic, but, the model’s average precision under the precision-recall curve actually increases during this time. This boost in precision, however, is not a reflection of increased discriminative ability between admission or discharge for the model, but actually the result of a change in the target variable distribution: the average patient attending in the COVID-19 pandemic is far more likely to be admitted (panel b in Figure 2) during this time period. Since precision only considers positive predictions (i.e. predicted admissions), a distribution skewed towards admissions could lead to a misleading performance metric. Evaluating the confusion matrices for pre and during COVID-19 at a fixed recall (0.75), shows that a degradation of performance would only be seen for patients who were actually discharged. During COVID-19 the lower model performance therefore appears as a higher ‘false-alarm’ rate (i.e. the model predicts a patient should be admitted when they are discharged).

**Figure 4.**
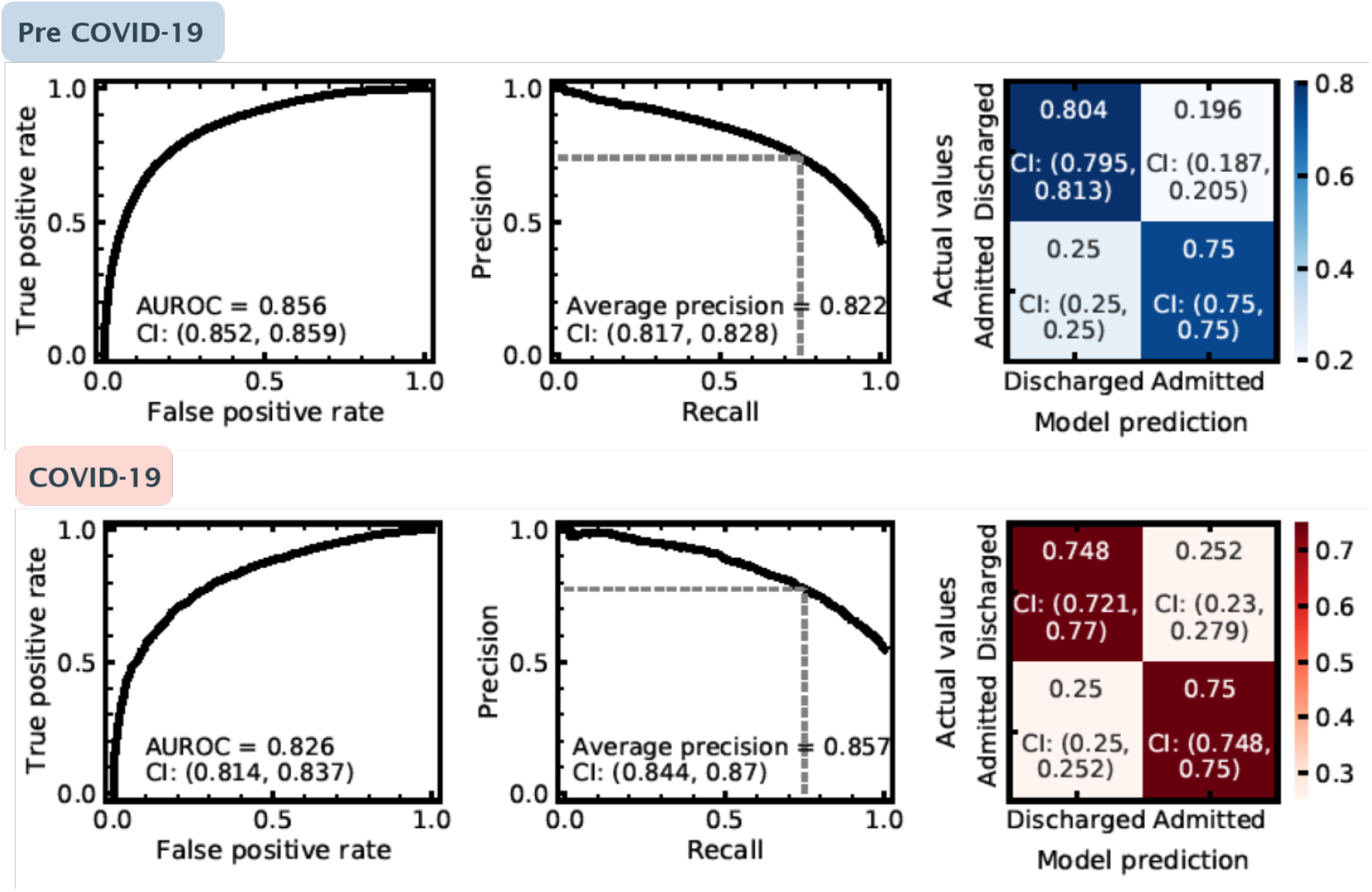
Performance of our admissions model evaluated pre COVID-19 (blue, top row) and during COVID-19 (red, bottom row). Panels left to right represent: 1) Receiver operating characteristic curve for the model’s predictions, 2) Precision recall curve for predictions on the test set, the dashed grey line shows the configuration evaluated in the confusion matrix in panel c., and c) Confusion matrix for admission predictions. Each panel shows the 2.5%, 97.5% confidence intervals on presented values, evaluated using bootstrapping (see main text).

In a medical context, it is also important to understand and track why a model makes its predictions, both for assurance purposes and to maximise the model’s functionality. Data drift is an important consideration in this domain. Fundamental changes to patient attendance behaviour should be tracked, regardless of how this presents in metrics such as the AUROC or average precision. Tracking how a model uses the information provided to it over time, enables a complimentary assessment of whether a model may need retraining.

Explainable machine learning enables attribution of the most important input features per model prediction. For our admissions model, this surmounts to having importance (SHAP) values defined for every feature, per attendance. These values can therefore be monitored periodically as a complimentary metric of detecting changes to demand, and operational policies. In the left panel of 5, we show the normalised SHAP values, binned weekly across our complete test dataset, of the top three features which increased in relative importance following the onset of COVID-19. In the right panel we show the same for the top three features which decreased in relative importance. Since the admission rate changes over time, so will the average sum across all features (see Model Explainability section for more discussion). We compute normalised SHAP values to ensure a consistent comparison of relative importance for a given feature.

For respiration rate, triage discriminator, and arrival mode we see a clear boost in average importance prior to and during the first UK national lockdown (23rd March onwards). While both respiration rate and triage discriminator decrease in their average importance post April 2020, arrival mode remains consistently more important than pre-COVID-19. Notably, respiration rate and triage discriminator are likely reflective of dynamically changing ED operations, and, highlight procedural change (or record-keeping) as the COVID-19 pandemic progressed. Conversely, arrival mode (to ED) may be reflective of a sustained change in patient behaviour throughout the first wave of the COVID-19 pandemic. As seen in Figure 2 (panel e) we found a significant decrease in patients attending through ‘walk-ins’, which leads to increased SHAP values for the feature because other modes of arrivals (e.g., ambulance arrivals) are generally used by patients who are more severely ill and are therefore more likely to be admitted. On-the-fly model evaluation (i.e. through a performance metric such as AUROC), along with normalised SHAP values would not only indicate a degradation in performance, but also, attribution of what is changing (e.g. features associated with clinical procedure or patient attendance behaviour). Tracking of normalised SHAP values could help highlight data drift as a trigger for model retraining before it may actually lead to significant performance degradation.

A specific example of how SHAP feature tracking could be used, is to identify emergent health risks. Respiration rate (Figure 5 top left panel), shows peaks both beginning in December 2019 (i.e. potentially reflecting the flu season), and March 2020 (i.e. onset of COVID-19). The weekly importance of respiration rate could therefore be used as an early indicator for the prevalence of viral complaints in the community; enabling hospitals to plan accordingly (e.g. if flu season peaks early). Conversely, SHAP feature tracking can provide early warnings of undesirable operational change. For example, if recorded vital signs drop in importance (with no change to the acuity of patients) it could be an indication of strain on time/resources with clinicians unable to fulfill comprehensive triage. Attribution of change in importance of attendance characteristics immediately point to changes in patient population, and operational change and could therefore enable early warning systems of emergent health risks for the regional population.

**Figure 5.**
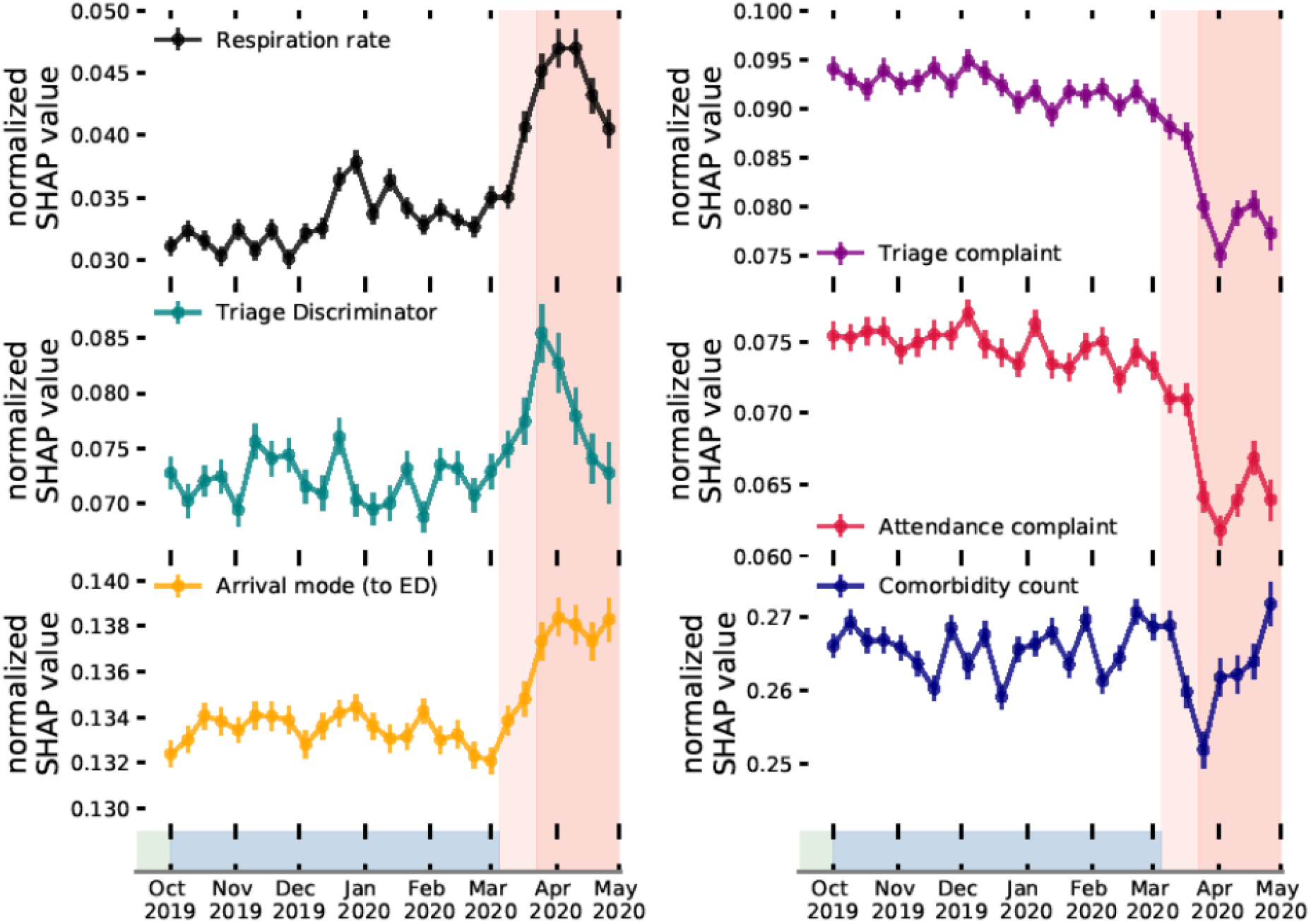
Time evolution of normalised SHAP values for most (left) and least (right) important features in COVID-19. (Top to bottom) normalised absolute SHAP values (a higher value shows the feature was proportionally more predictive at the given time) of respiration rate, triage discriminator, and, arrival mode (left). Values for triage complaint, attendance complaint, and, comorbidity count (right). The red shading is consistent with the timeline in Figure 1.

### Data drift in COVID-19

We now discuss the use of explainable machine learning to further characterise the data drift observed as a result of the COVID-19 pandemic and provide specifics of how it impacted operational circumstances for EDs. In Figure 6, we consider which input features were most important for our admissions model. In the left panel, we show the overall feature importance ranking to understand the most predictive variables in determining admission risk. This is found by the mean absolute SHAP value across both pre and during COVID-19 patient cohorts. The most important variables include the number of comorbidities, arrival mode, and patient age which all latently encode a patient’s fraility; highly predictive of admission risk^19^. In addition, features indicating the patient’s reason for attendance (attendance and triage complaints, triage discriminator) are also strongly predictive of admission risk.

**Figure 6.**
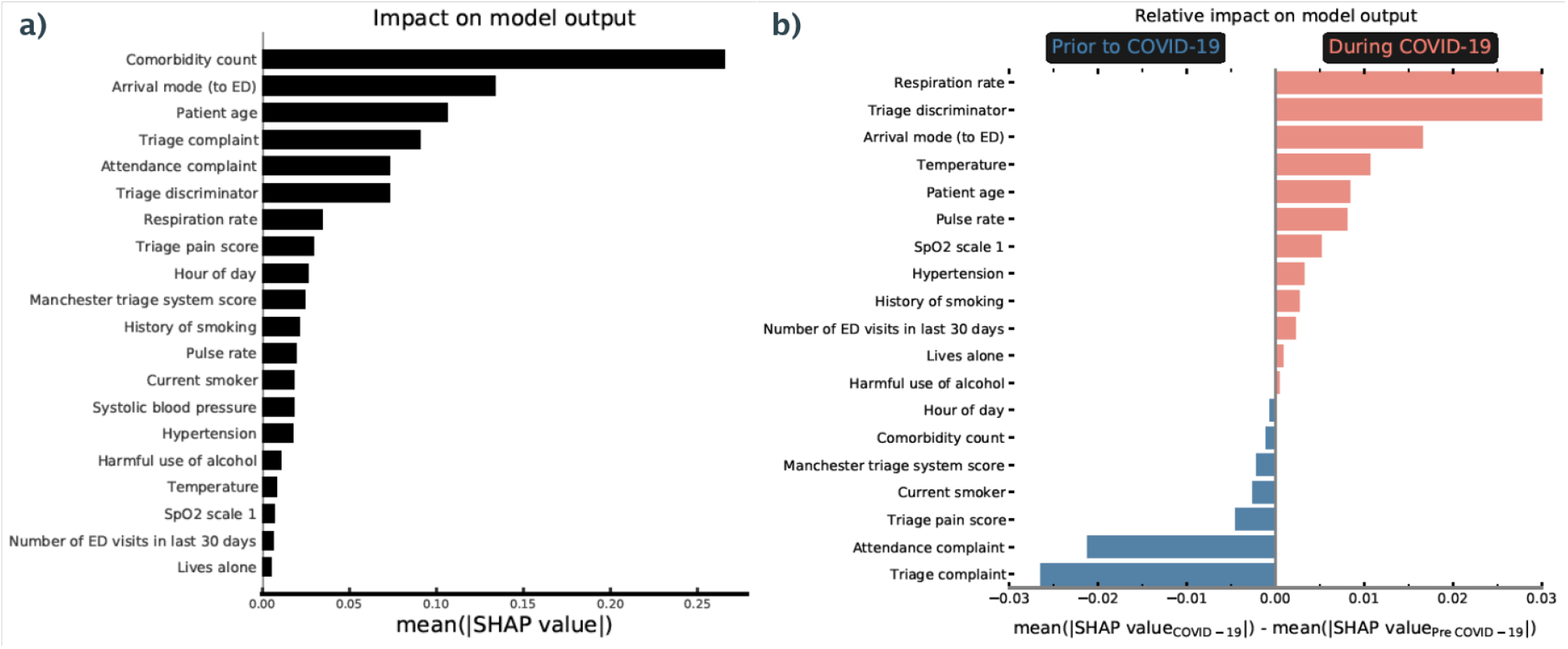
Importance ranking of input features for predicting admission risk. a) Average (absolute) SHAP value for attendance features over all patients (both pre and during COVID). A higher value corresponds to a more important feature in decision making (i.e. predictive of admission risk). b) Relative importance ranking of input features for predicting admission risk in COVID-19. Average (absolute) SHAP value for attendance features during COVID-19 relative to pre COVID-19 values. A higher (lower) value corresponds to a more important feature in decision making during (pre) COVID-19.

In the right panel of Figure 6 we show which features changed in importance for predicting admission risk during the COVID-19 pandemic. Here we find the mean absolute SHAP values for pre and during COVID-19 patient cohorts respectively and find the difference between the two (where positive values show more important features during COVID-19). One key theme is the increased importance of recorded vital signs in COVID-19 (e.g. respiration rate, temperature, pulse rate, SpO2 scale). A practical explanation for the increased importance of these features could simply be the rate at which they are recorded. Panel c) of Figure 2 shows that the fraction of patients with recorded vital signs increases sharply during COVID-19, potentially indicative of clinician’s increased dependence on vital signs for risk assessment. Another possibility is that the average severity of complaint during the COVID-19 pandemic increased (e.g. patient physiological change so that minor injuries were no longer attending) which itself leads to a higher rate at which patient’s vital signs are taken.

The frequency of missing values are also important point in proactively monitoring machine learning models. A change in record keeping could indicate significant procedural change, and, in doing so lead to instability in performance. During the COVID-19 pandemic additional complaint categories (e.g. ‘COVID-19’) were eventually added to hospital record keeping systems. For a model trained pre-COVID-19, any patient designated under ‘COVID-19’ would be treated as missing information for the category, which the model would not know how to optimally use. SHAP values immediately highlight which recorded features become significantly less useful, therefore enabling automatic detection of procedural change, or changes to clinical reporting behaviours.

Another significant shift is the increased importance (during COVID-19) of ‘triage discriminator’ and the decrease in ‘attendance (and triage) complaint’. The complaints describe the overall reason for patient attendance at the point of registration (attendance) and triage, whereas triage discriminator includes additional sub-categories about the complaint. To understand these shifts, in Figure 7 we show the three most prevalent categories during (and pre) COVID-19 for attendance complaint, triage complaint, and, triage discriminator. Generic and uninformative categories are significantly more prevalent during COVID-19 for the complaints (e.g. unwell adult), whereas specific problems such as head injuries and wounds are far less prevalent. Conversely for triage discriminator (right panel), specific categories such as pleuritic pain and (very) Low SpO2 directly related to viral infections become far more prevalent, whereas uninformative categories (e.g. recent problem, recent mild pain) were far more common pre COVID-19. These changes in categorical prevalence intuit why a given feature became more or less predictive of admission risk during the COVID-19 pandemic. In particular, we note that the most prevalent categories (during COVID-19) in triage discriminator likely encode symptomatic COVID-19. With no categorical variable attributed to COVID-19, clinical record-keeping instead reflected vague or unspecific complaints while identifying specific viral symptoms in the discriminator.

**Figure 7.**
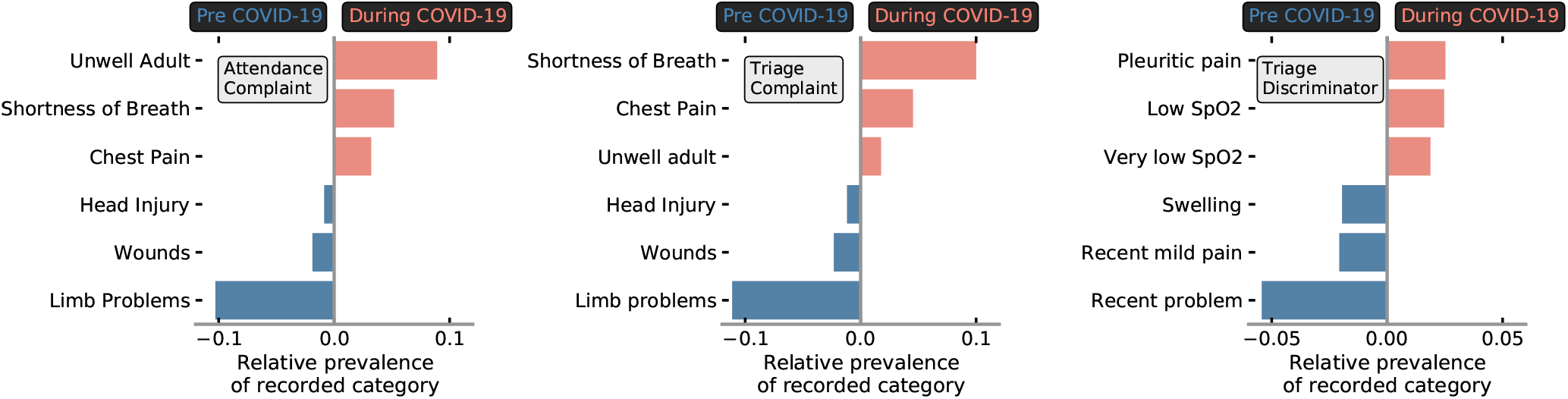
Relative prevalence of patient records (i.e. that indicate a patient’s symptoms) in COVID-19. Average prevalence of recorded categories during COVID-19 (relative to pre COVID-19) recorded under attendance complaint (left), triage complaint (middle), and, triage discriminator (right). A higher (lower) value corresponds to a more prevalent complaint during (pre) COVID.

## Discussion

In this work we have demonstrated the potential for explainable machine learning to be used as a complimentary approach to detect and understand data drift for models deployed in dynamic environments, such as EDs. We have trained and retrospectively evaluated a machine learning classifier which predicts the risk of a patient of being admitted to hospital following an ED attendance. Using features describing the ED attendance (e.g. basic patient information and condition history, attendance information and chief complaint), our model achieved an AUROC of 0.856 (top left in Figure 4) on a (pre-COVID-19) test set. This represents state-of-the-art performance comparable with recent admission models (e.g. AUROC 0.825-0.877^20–23^).

Using the COVID-19 as an exemplar for data drift, we introduce the severe changes to attendance features in EDs during the COVID-19 pandemic (Figure 2), summarised by a large drop off in patient attendance (driven by a lack of ‘walk-ins’), a sharp increase in admission rate, a reduction in the number patients attending for wounds and traumas, and an increase in patients presenting with viral complaints. We demonstrate that this significant circumstantial change during the first wave of the pandemic is correlated with a small degradation of model performance (AUROC = 0.83; bottom left in Figure 4). Due to a change in the target distribution, (i.e. significantly more patients being admitted to hospital) precision, perhaps misleadingly, increases. Using performance metrics alone, it is difficult to ascertain whether the change is resultant from a shift in the underlying statistical relationship between features and target variables (concept drift), or a shift in the covariates to be more closely distributed to regions where the model’s local performance is lower (data drift). In a medical context, data drift (even with no degradation in model performance) is important to detect, as changes to patient behaviour or ED operations should be continually assessed to understand the factors which could drive changes in model output. Commonly adopted techniques for tracking data drift include univariate statistical tests^24,25^ (e.g. Kolmogorov-Smirnov tests to distinguish changes in data distributions) and adversarial validation^26,27^ (i.e. training external models to detect different data domains). However, these act externally to the classifier, and tracking how a model uses the information provided to it over time, could enable a complimentary assessment of whether a model needs retraining.

Using explainable machine learning, we are able to determine the importance of input features, per attendance, enabling identification of data drift. We track the importance of given model features on a weekly basis (Figure 5) through normalised SHAP values. The importance attribution of normalised SHAP values give the method a clear advantage over directly monitoring the distributions of input features. SHAP directly quantifies how impactful the data drift has been per covariate for the model, rather than only assessing the magnitude of change, and hence, provides more insight into whether a model requires retraining. Used in combination with performance metric tracking, importance monitoring represents a powerful methodology of both determining when a model may require retraining, and attributing why model performance degradation may be occurring. In addition, if model performance deteriorates in absence of statistically significant change to the normalised SHAP values, it is directly indicative of concept drift. In this instance stable SHAP values indicates no change to model decision making, and hence, a deterioration in performance points to a change in the relationship of features to target variables. This aspect is particularly useful for EDs since it directly highlights changes to admission decision making, perhaps under capacity pressures (which could be used to allocate more staff). Monitoring importance is critical in ensuring any deployed model, and ED remains fair and unbiased in assessing each patient’s risk. We also note the potential for SHAP to be used to identify emergent diseases. Using the example of respiration rate (Figure 5 top left panel) we find that this feature both tracks the onset of flu season (Dec 2019) and COVID-19 (March 2020). Feature tracking could therefore act as an early warning system of diseases emerging in the regional patient population.

For our model, SHAP values can also be used to determine which features are of most importance for overall admission risk, and to contrast what changed during the COVID-19 pandemic. Overall, the number of comorbidities, arrival mode, and patient age are highly predictive of admission risk (Figure 6 panel a). These are all factors that latently encode patient frailty, closely related to both higher long-term admission and re-admission risk^28,29^. To understand what features became more (or less) important for our to model to determine admission risk during COVID-19, we used the relative difference between average SHAP values for pre and during COVID-19 patient cohorts (Figure 6 panel b). We find that vital signs (e.g. respiration rate, pulse rate, SpO2 levels), arrival mode, and, triage discriminator became more important for model predictions of patient outcomes during COVID-19. Due to the significant drop of patients attending through ‘walk-ins’ (Figure 2; typically associated with less frail attendances), arrival mode likely became more important as the average severity of attendances increased. Conversely triage discriminator, and, vital signs are reflective of procedural change. Within UHS, a combination of vital signs were used to assess COVID-19 risk. This led to sharp up-tick in the fraction of patients with recorded vital signs (panel c, Figure 2), picked up by a boost in importance in our model explainablity. We find that triage discriminator *latently encoded* symptomatic COVID-19 (Figure 7) through higher prevalence of respiratory related categories. Attendance/triage complaint more frequently contained uninformative categories (i.e. ‘unwell adult’) due to a lack of a COVID-19 category (not added to the ED’s record keeping system in the initial stages of the COVID-19 pandemic).

A possible implementation of an ED admission prediction model, is ahead-of-time resource management and logistical planning both within the ED itself, and, in downstream care. Identifying the requirement of bed space (or other resources) for a patient at the first point of contact (either at ambulance pick-up or patient registration in the ED) enables hospitals to pre-allocate resources on acuity. Admission risk appears to be largely dictated by patient frailty and attendance complaint, and, hence admission models implemented early in the ED visit pathway are still highly predictive. We chose to implement our model at the point of triage, since ahead-of-time logistical management both within the ED and throughout the patient’s care trajectory is still possible, while aiding model explainability which concerns operational procedure (i.e. including vital sign records and triage discriminator as features). One limitation of this study is that we only possess prior patient information from previous discharge summaries at UHS, and, hence only have patient histories for 55% of our recorded attendances. Including patient histories from their non-emergency and routine treatment would enable even more predictive early warning modelling.

## Conclusion

In conclusion, we have trained a machine learning classifier to identify patients at high risk of admission to hospital at the point of attendance to an ED. On a hold-out pre-COVID-19 test set our classifier obtained an AUROC of 0.856 95%CI [0.852, 0.859]. Using the COVID-19 pandemic as an exemplar of data drift, we evaluated our model’s performance during severe operational change with our classifier obtaining an AUROC of 0.826 95%CI [0.814, 0.837]. We demonstrated how explainable machine learning can be used attribute changes in feature importance over time, as a complimentary metric to detect data drift, and emergent health risks. One limitation of this work is that we cannot formally detect concept drift (only data drift). In future work we look to formally combine explainable machine learning with various techniques to isolate concept drift.

## Data Availability

The data that support the findings of this study are available from UHS, but restrictions apply to the availability of these data, which were used under license for the current study, and so are not publicly available. Data are however available from the authors upon reasonable request and with permission of UHS.

## Acknowledgements

This work was fully supported by The Alan Turing Institute. We acknowledge Martin Azor and Florina Borca for data extraction support. We thank Sarah Reeves for additional clinical insight.

## Author contributions statement

CJD and FPC performed the data analysis and modelling. ZDZ, DKB, and MJB discussed and commented on the analysis with CJD. NW and FPC obtained governance and ethical approval. CJD, FPC, MJB and NW managed the study at the UoS. MK managed the study at UHS. CJD, FPC and MK designed the study with assistance from TWVD. MK and TWVD provided clinical guidance and insight. CJD wrote the first draft of the manuscript. All authors frequently discussed the work and commented and contributed to future drafts of the manuscript.

## Additional information

### Data availability

Due to patient privacy concerns the dataset used in this study is not publicly available. However, it will be made available upon reasonable request.

