## Supplementary Table 1 for "Emergency department admissions during COVID-19: explainable machine learning to characterise data drift and detect emergent health risks"

### Supplementary Material

| Name | Encoding | Additional notes |
|---|---|---|
| 30 day visit count | N/A | ED visits by patient in last 30 days |
| Hour of day | N/A | |
| Weekday | N/A | |
| Patient age | N/A | |
| Triage category | Target encoding | Manchester triage system score |
| Triage painscore | N/A | Alder Hey triage pain score |
| Respiration rate | N/A | Breaths per minute |
| SpO2 scale 1 | N/A | Pulse oximetry (oxygen saturation) |
| Systolic blood pressure | N/A | |
| Pulse rate | N/A | |
| Temperature | N/A | |
| History of smoking | One-hot encoding | |
| Current smoker | One-hot encoding | |
| Hypertension | One-hot encoding | |
| Harmful use of alcohol | One-hot encoding | |
| Depression | One-hot encoding | |
| Type 2 diabetes | One-hot encoding | |
| Asthma | One-hot encoding | |
| Lives alone | One-hot encoding | |
| COPD | One-hot encoding | |
| Alcohol screening | One-hot encoding | |
| Harmful use of alcohol with dependence | One-hot encoding | |
| Osteoarthritis | One-hot encoding | |
| Hypercholesterolaemia | One-hot encoding | |
| Anxiety | One-hot encoding | |
| Atrial fibrillation | One-hot encoding | |
| Anaemia | One-hot encoding | |
| Obesity | One-hot encoding | |
| Chronic kidney disease | One-hot encoding | |
| Hypothyroidism | One-hot encoding | |
| Type 1 diabetes | One-hot encoding | |
| Number of comorbidities | N/A | Comorbidities for patient listed on previous ED visits. |
| Arrival mode | Target encoding | |
| Attendance complaint | Target encoding | Chief complaint taken at attendance |
| Triage complaint | Target encoding | Chief complaint taken at triage |
| Triage discriminator | Target encoding | Complaint additional notes taken at triage |

**Table 1.** Full list of features used as input for the admissions model. For each feature, the type of encoding (if applicable) is noted along with any additional explanatory remarks. The one-hot encoded comorbidities are limited to the top 20 most common, however, the number of comorbidities counts all listed in the UHS database.
